# Angiographic Tool to Detect Pulmonary Arteriovenous Malformations in Single Ventricle Physiology

**DOI:** 10.1101/2024.01.08.24300994

**Authors:** Stephen B Spurgin, Yousef M Arar, Thomas M Zellers, Jijia Wang, Nicolas L Madsen, Surendranath R Veeram Reddy, Ondine Cleaver, Abhay A Divekar

**Affiliations:** Department of Pediatrics, Southwestern Medical Center, Dallas, TX 75390, USA; Pediatric Cardiology, Children’s Medical Center, Dallas, TX 75235, USA; Department of Applied Clinical Research, University of Texas Southwestern Medical Center, Dallas, TX 75390, USA; Department of Molecular Biology and Center for Regenerative Science and Medicine, University of Texas Southwestern Medical Center, Dallas, TX 75390, USA

**Keywords:** Pulmonary arteriovenous malformation, superior cavopulmonary anastomosis, pulmonary transit time, hepatic factor

## Abstract

**Background:** Individuals with single ventricle physiology who are palliated with superior cavopulmonary anastomosis (Glenn surgery) may develop pulmonary arteriovenous malformations (PAVMs). The traditional tools for PAVM diagnosis are often of limited diagnostic utility in this patient population. We sought to measure the pulmonary capillary transit time (PCTT) to determine its value as a tool to identify PAVMs in patients with single ventricle physiology.

**Methods:** We defined the angiographic PCTT as the number of cardiac cycles required for transit of contrast from the distal pulmonary arteries to the pulmonary veins. Patients were retrospectively recruited from a single quaternary North American pediatric center, and angiographic and clinical data was reviewed. PCTT was calculated in 20 control patients and compared to 20 single ventricle patients at the pre-Glenn, Glenn, and Fontan surgical stages (which were compared with a linear-mixed model). Correlation (Pearson) between PCTT and hemodynamic and injection parameters was assessed using 84 Glenn angiograms. Five independent observers calculated PCTT to measure reproducibility (intra-class correlation coefficient).

**Results:** Mean PCTT was 3.3 cardiac cycles in the control population, and 3.5, 2.4, and 3.5 in the pre-Glenn, Glenn, and Fontan stages, respectively. PCTT in the Glenn population did not correlate with injection conditions. Intraclass correlation coefficient was 0.87.

**Conclusions:** Pulmonary angiography can be used to calculate the pulmonary capillary transit time, which is reproducible between observers. PCTT accelerates in the Glenn stage, correlating with absence of direct hepatopulmonary venous flow.

## INTRODUCTION

Pulmonary arteriovenous malformations (PAVMs) frequently develop after superior cavopulmonary anastomosis (Glenn surgery) (1-4). Remarkably, these PAVMs can resolve after total cavopulmonary anastomosis (Fontan completion), which has led to the “hepatic factor” hypothesis: the liver makes an unknown substance that maintains the normal vascular architecture of the lungs (3, 5). Surgical and interventional strategies to redirect hepatic venous blood to affected lungs have been successful to resolve PAVMs (6-9). However, while all Glenn patients lose direct flow of “hepatic factor” to their lungs, not all Glenn patients develop clinically significant PAVMs (10).

PAVM screening of Glenn patients is fraught with technical difficulty. Intracardiac mixing prevents use of peripheral arterial PaO_2_ saturation. Direct measurement of PaO_2_ from pulmonary veins is not always feasible. The common screening modality (peripherally injected bubble contrast echocardiography) is technically variable, prone to false positives, and unable to detect capillary recruitment or proliferation of small caliber vessels (11-14). Meanwhile, the confirmatory test used in patients with hereditary hemorrhagic telangiectasia (computed tomography scan) has a spatial resolution of 0.15 mm, which is 10 times greater than the size of the dilated capillary vessels that could allow for positive contrast echocardiogram (14, 15). Therefore, CT may not be able to detect shunts that are due to diffuse microvascular dilation.

In light of these unresolved issues with the current tools for detection, and cognizant of the importance of detection for prognosis and clinical management, we sought to develop a screening tool that could detect changes in the pulmonary microvascular architecture. We used pulmonary angiography to define a pulmonary capillary transit time (PCTT), tracking the flow of contrast through the normal pulmonary vasculature—or through the shortcuts that PAVMs provide. This PCTT tool, applied to single ventricle patients with Glenn anatomy, was studied to determine pulmonary vascular changes that may occur in the absence of direct hepatopulmonary venous flow.

## MATERIALS and METHODS

### Study Cohort

Single ventricle patients who underwent clinically indicated cardiac catheterization including a pulmonary artery angiogram at our institution between February 2021 and February 2023 were included in the study. For these patients, both current and prior angiograms were reviewed. Baseline clinical and catheterization data (hemodynamic measurements, blood gases, and injection parameters) for all patients was analyzed, in accordance with UTSW IRB 2020-0047.

A group of twenty patients with biventricular circulation and normal hepatopulmonary blood flow served as controls (pulmonary artery stenosis, n=11; pulmonary valve stenosis, n=9). Twenty patients with univentricular hearts and pulmonary angiograms available from all three surgical stages were selected for longitudinal analysis.

Eighty-four Glenn-stage angiograms were utilized to evaluate for correlation between PCTT and injection conditions, hemodynamic measurements, or additional clinical factors. PCTTs for this group were calculated by the lead author. From this group, twenty sequentially-enrolled angiograms were evaluated by the lead author as well as four pediatric interventional cardiologists (YA, TZ, SR, AD) to measure the reproducibility of the visually-calculated PCTT.

### Definition of PCTT

We defined PCTT as the number of cardiac cycles between initial opacification of the distal pulmonary arteries to the earliest opacification of the major pulmonary veins, indexed to the cardiac cycle **(Figure 1**). The degree of opacification was not taken into account. All patients were sedated and mechanically ventilated as part of the procedure, and angiographic technique was at the discretion of the attending cardiologist. However, angiograms were only selected for analysis if the contrast was injected centrally, not in a distal pulmonary artery (**Supplementary Table S1**). Consequently, there is a short delay between the start of injection and the start of the PCTT.

**Figure 1:**
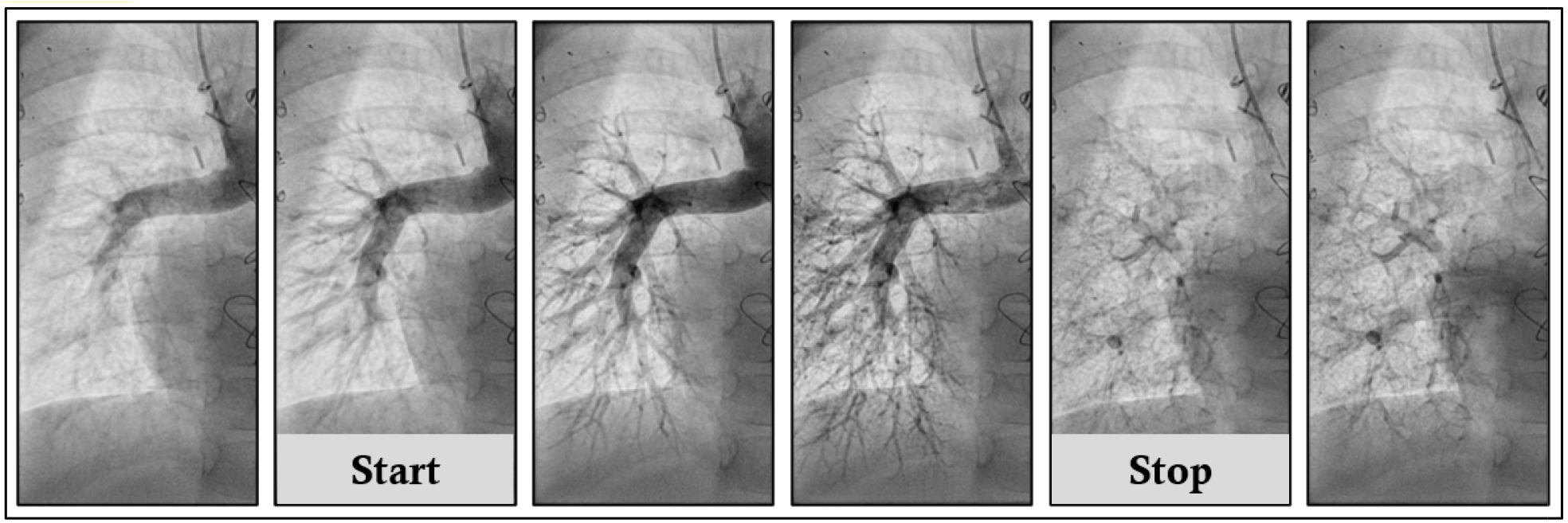
Definition of the PCTT. Pulmonary capillary transit time is defined as the number of cardiac cycles taken for contrast to traverse the pulmonary capillary bed. Calculation of the PCTT starts when the contrast reaches the distal major branches of the pulmonary arteries, and stops when the first visible contrast is present in the pulmonary veins.

Cardiac cycle time was obtained from simultaneous EKG recordings. When simultaneous EKG was not available, direct observation of the motion of the cardiac silhouette was used (average of five cardiac cycles; frame rate of the angiogram divided by number of frames per beat, multiplied by 60 to obtain beats per minute). PCTT was calculated for left and right lungs separately, and the average was then used for subsequent grouping and analysis.

### Statistical Analysis

Descriptive statistics (i.e., mean, standard deviation, median, min, and max) were employed for summarizing demographic, hemodynamic, and clinical variables. The two-sample t test was used to compare continuous variables. ANOVA was used to compare the continuous variables between several groups with Tukey HSD being the multiple comparison adjustment method. The relationships between PCTT and injection pressure, dose, rate, and rise were investigated by the Pearson correlation coefficient. For comparing the PCTT in the longitudinal analysis, a linear mixed model was used to incorporate the correlation between repeated measurements from the same patient. Intraclass correlation coefficient (ICC) based on the two-way random effects model was used to investigate the agreement between the five readers. The p values for pairwise ICC were not adjusted. The level of significance was set at 5%. All the analyses were conducted using SPSS v29 (IBM Corp., Armonk, NY), GraphPad Prism 10.0.0 (Dotmatics, La Jolla, CA), or SAS 9.4 (SAS Inc., Cary, NC).

## RESULTS

### PCTT is Accelerated During the Glenn Stage

The clinical characteristics of our Glenn cohort are listed in **Table 1**. We found that the mean PCTT of the control group (with normal cardiopulmonary vascular connections and biventricular hearts) was 3.25 (95% CI 2.9 – 3.6). Similar to the control group, the mean PCTT of the pre-Glenn and the Fontan groups were 3.53 and 3.48, respectively (**Figure 2A**). However, the mean PCTT of the Glenn patients was 2.37 (95% CI 2.01 – 2.73) cardiac cycles, significantly faster than all other individual groups (p<0.001). A detailed comparison of clinical characteristics at each stage is available in **Supplementary Table S2**.

**Table 1:** Study Cohort Characteristics. Among 84 Glenn patients recruited to the study, the majority were male, situs solitus, with a single right ventricle and variable initial palliation procedures.

|  |  | N (%) |
| --- | --- | --- |
| Sex | Male | 54 (64) |
|  | Female | 30 (36) |
| Functional Ventricle | Right | 60 (71.4) |
|  | Left | 24 (28.6) |
| Major Cardiac Defect | HLHS | 33 (39.3) |
|  | DORV | 16 (19.0) |
|  | AVCD | 11 (13.1) |
|  | TA | 10 (11.9) |
|  | PA | 8 (9.5) |
|  | DILV | 6 (7.1) |
| Visceral Situs | Solitus | 69 (82.1) |
|  | Inversus | 1 (1.2) |
|  | Right Isomerism | 6 (7.1) |
|  | Left Isomerism | 8 (9.5) |
| Initial Palliation | Sano | 28 (33.3) |
|  | BTTS | 21 (25.0) |
|  | PDA Stent | 13 (15.5) |
|  | PA Band | 11 (13.1) |
|  | None | 11 (13.1) |
| Glenn Sidedness | Right | 67 (79.8) |
|  | Left | 9 (10.7) |
|  | Bilateral | 8 (9.5) |
| Antegrade Pulmonary Blood Flow | No | 78 (92.9) |
|  | Yes | 6 (7.1) |
| PA Plasty at Time of Glenn | No | 40 (47.6) |
|  | Yes | 44 (52.4) |

**Figure 2:**
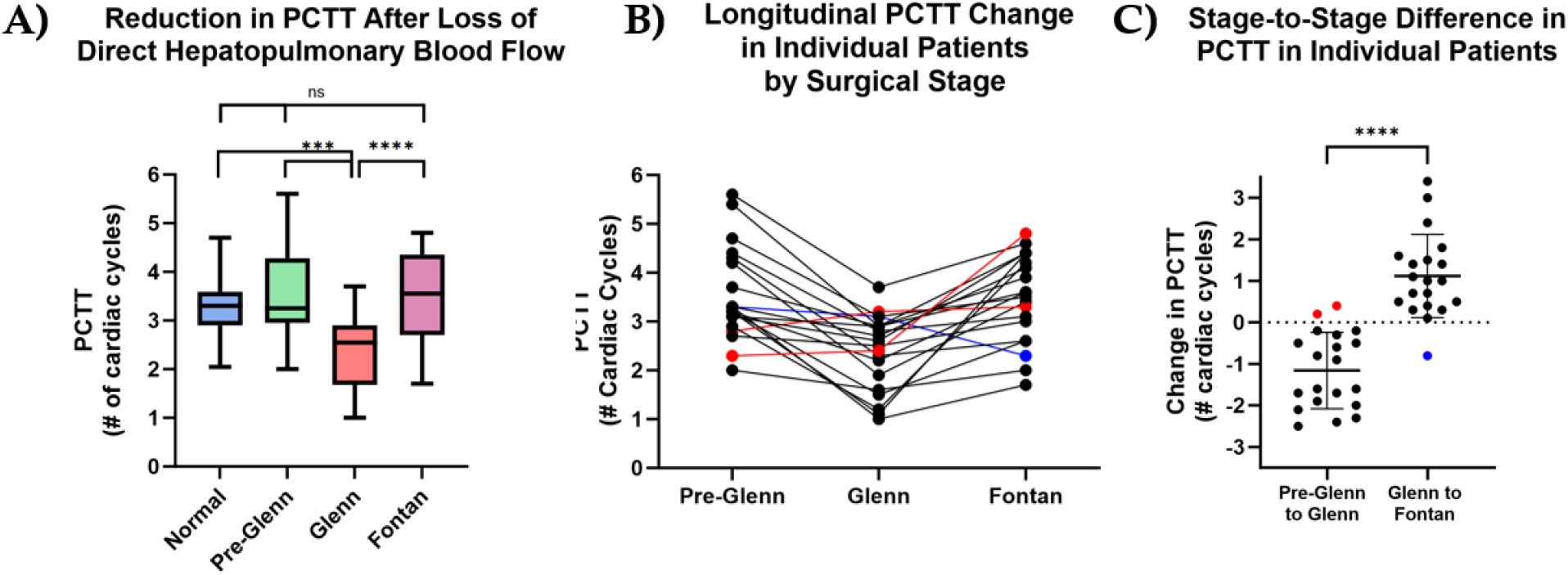
Loss of Direct Hepatopulmonary Blood Flow Leads to Reversible Acceleration of PCTT. **(A)** There is no significant difference in the PCTT between patients with normal cardiopulmonary vascular connections and single ventricle patients in the Pre-Glenn or Fontan stage. However, the PCTT in patients with Glenn anatomy is significantly accelerated. **(B)** Tracking individual patients (from A) through each surgical stage shows accelerated PCTT in the Glenn that reverts to normal after restoration of direct hepatopulmonary blood flow. Two patients whose PCTT did not decrease from Pre-Glenn to Glenn are highlighted in red, and one patient whose PCTT did not increase from Glenn to Fontan is highlighted in blue. **(C)** Stage-to-stage difference for the individual patients shown in (B).

When individual patients were analyzed longitudinally, we found that 90% (18/20) of patients had a faster PCTT in the Glenn stage, compared to the pre-Glenn stage (**Figure 2B,C**). Using a linear mixed model, incorporating the correlation between repeated measurements from the same patient, we found that there is a significant decrease from pre-Glenn to Glenn (p < 0.0001) and a significant increase from Glenn to Fontan (p = 0.0002) (**Table 2**).

**Table 2:**
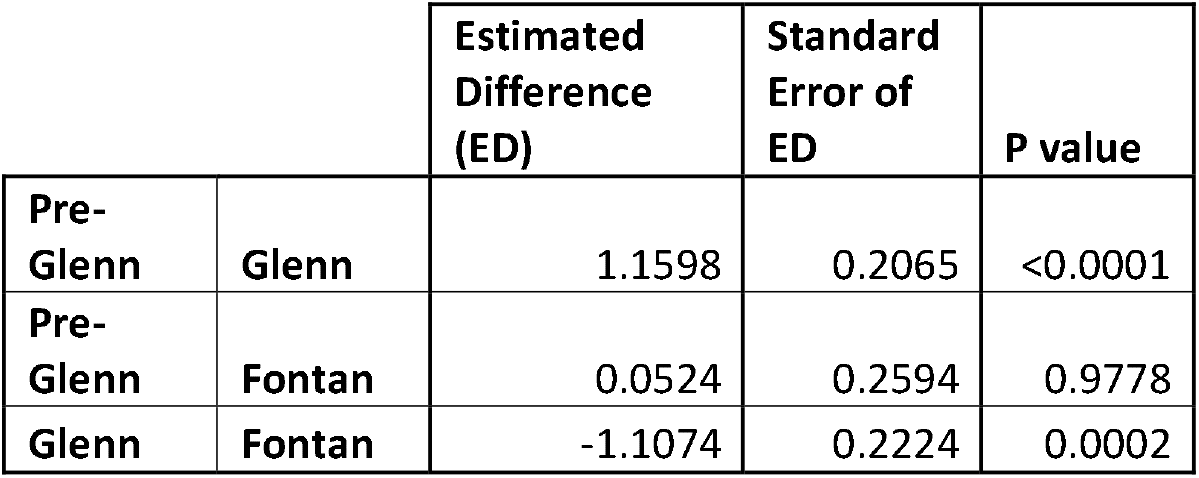

|  |  | Estimated Difference (ED) | Standard Error of ED | P value |
| --- | --- | --- | --- | --- |
| Pre-Glenn | Glenn | 1.1598 | 0.2065 | <0.0001 |
| Pre-Glenn | Fontan | 0.0524 | 0.2594 | 0.9778 |
| Glenn | Fontan | -1.1074 | 0.2224 | 0.0002 |

In our Glenn patients, there was no association between PCTT and either injection pressure, contrast dose, rate of injection, or rate of rise (**Supplementary Figure S1**). Additionally, there was no association between PCTT and catheter size (p=.55) or catheter type (p=.24) (**Supplementary Figure S2**). Similarly, in the Glenn cohort, neither peak PA pressure, mean PA pressure, transpulmonary gradient, or ventricular end-diastolic pressure correlated with PCTT (**Table 3**). Five independent reviewers of calculated PCTT for twenty Glenn angiograms, which yielded an ICC of 0.87 (95% CI 0.7 – 0.95, p<0.001), indicating good agreement between viewers (**Supplementary Table S3**).

**Table 3:** Relation of PCTT and Patient-Specific Factors During the Glenn Stage. Basic physical parameters, angiographic and hemodynamic conditions, and readouts of successful alveolar gas exchange were examined using Pearson correlation for their relation to the PCTT. Parameters were highlighted in red if there was a significant correlation with PCTT. *All pressure measurements are in mm Hg. Abbreviations: PA=pulmonary artery, PV=pulmonary vein, PCW=pulmonary capillary wedge, Qp=pulmonary blood flow, Qs=systemic blood flow, PVR=pulmonary vascular resistance (Woods units), SVR=systemic vascular resistance (Woods units), RUPV=right upper PV, RLPV=right lower PV, LUPV=left upper PV, LLPV=left lower PV*.

|  |  | Mean Value |  | Correlation With PCTT |  |
| --- | --- | --- | --- | --- | --- |
| | | N | Mean ± SD | r | P value |
| Demographics | Age (Years) | 84 | 3.8 ± 1.5 | 0.026 | 0.816 |
| | Age at Glenn | 84 | 0.6 ± 0.6 | -0.16 | 0.141 |
| | Time Since Glenn | 84 | 3.2 ± 1.5 | 0.094 | 0.396 |
| | Age at Fontan | 47 | 4.5 ± 1.3 | -0.074 | 0.619 |
| | Time Spent as Glenn | 47 | 4.0 ± 1.3 | -0.00035 | 0.998 |
| Physical Parameters | Height (cm) | 84 | 94.4 ± 17.5 | 0.1 | 0.354 |
| | Weight (kg) | 84 | 15.9 ± 10.5 | 0.013 | 0.905 |
| | BSA (m<sup>2</sup>) | 84 | 0.6 ± 0.2 | 0.062 | 0.577 |
| | VO2 (mL/kg/min) | 84 | 159.5 ± 26.1 | 0.042 | 0.703 |
| Catheterization Hemodynamics | Heart Rate | 84 | 88.8 ± 14.9 | 0.19 | 0.091 |
| | Peak PA Pressure | 84 | 13.5 ± 2.7 | -0.11 | 0.308 |
| | Trough PA pressure | 84 | 12.4 ± 2.5 | -0.027 | 0.808 |
| | Delta PA Pressure | 84 | 1.2 ± 1.3 | -0.18 | 0.096 |
| | Mean PA pressure | 84 | 12.7 ± 2.4 | -0.078 | 0.480 |
| | Mean PV or PCW Pressure | 84 | 8.9 ± 1.8 | -0.11 | 0.318 |
| | TPG | 84 | 3.8 ± 1.5 | 0.0087 | 0.937 |
| | Ventricular EDP | 84 | 9.0 ± 2.0 | -0.085 | 0.440 |
| Fick Calculation of Flows/Resistances | Qp (mL/min) | 73 | 2.7 ± 0.7 | -0.16 | 0.186 |
| | Qs (mL/min) | 77 | 4.5 ± 2.5 | -0.053 | 0.648 |
| | Qp:Qs | 73 | 0.6 ± 0.2 | 0.035 | 0.768 |
| | PVR (Woods units) | 71 | 1.7 ± 0.6 | 0.16 | 0.177 |
| | SVR (Woods units) | 72 | 12.6 ± 4.8 | 0.09 | 0.450 |
| PV O2 Sat | Minimum PV Sat | 68 | 94.4 ± 7.4 | 0.37 | 0.002 |
| Femoral Artery Blood Gas | Femoral artery pH | 84 | 7.3 ± 0.0 | 0.063 | 0.570 |
| | PCO2 | 84 | 41.7 ± 5.0 | -0.016 | 0.882 |
| | PO2 | 84 | 52.6 ± 6.9 | 0.21 | 0.054 |
| | O2 sat | 84 | 83.7 ± 5.9 | 0.24 | 0.028 |
| | Base Excess | 84 | -3.8 ± 3.2 | 0.06 | 0.586 |
| | HCO3 (mEq/L) | 84 | 22.1 ± 2.8 | 0.06 | 0.588 |
| | Hemoglobin (g/dL) | 84 | 14.8 ± 1.8 | 0.011 | 0.918 |
| Contrast Injection Parameters | Catheter Size (French) | 84 | 5 ± 0.6 | -0.032 | 0.771 |
| | Dose (mL) | 84 | 11.1 ± 3.3 | -0.065 | 0.557 |
| | Rate (mL/sec) | 69 | 13.7 ± 2.8 | -0.17 | 0.156 |
| | Rise (sec) | 69 | 0.1 ± 0.3 | -0.029 | 0.811 |
| | Pressure (PSI) | 69 | 430.8 ± 162.6 | -0.18 | 0.145 |

## DISCUSSION

This study provides the first published evidence that the pulmonary capillary transit time varies with the stage of single ventricle palliation. Our results validate the traditional teaching that, in the absence of PAVMs, contrast takes at least three cardiac cycles to travel through the pulmonary capillary bed. In 90% (n=20) of patients, PCTT accelerates in the Glenn stage. The pre-Glenn data presented in our study provides evidence that the pulmonary vasculature was not initially abnormal (with a fast PCTT). Rather, the acceleration of pulmonary capillary transit correlates with the absence of direct hepatopulmonary venous flow in this surgical stage.

PAVM detection is difficult and subject to variable limitations in the Glenn population. Diagnostic modalities of PAVM detection may be understood as attempts either to *visualize the lesion* (CT, magnetic resonance imaging, MRI, or pulmonary angiography), or to *detect the effects* of pulmonary arteriovenous shunting (pulmonary venous desaturation, contrast echocardiography [CE], 99mTc macroaggregated albumin scan). The PCTT represents a combination of these two strategies, as we visualize the effects of pulmonary arteriovenous shunting.

Peripherally-injected contrast echocardiography (CE) remains the standard screening method for PAVMs in the general population (15). CE has been used previously to show reversibility of PAVMs after reintroduction of direct hepatopulmonary venous flow, by comparing patients in the Glenn and Fontan stages (5). Recent work in Fontan patients, however, has shown that the significant majority of Fontan patients have a positive CE (12). While we suspect that their remarkably high positive rate is affected by the use of a distally placed end-hole catheter for bubble contrast injection (applying direct force to dilate the peripheral capillaries), their work does highlight the need for a new diagnostic modality that can track changes secondary to presence or absence of hepatic factor.

While our work is the first of its kind in pediatric cardiology, a single study in adult patients with hepatopulmonary syndrome defines a similar “pulmonary transit time” (PTT, time in seconds from initial injection and opacification of the main pulmonary artery to that of the left atrium) (16). Intriguingly, they report a diagnostic utility comparable to contrast echocardiography. However, they include in their transit time the portion that takes place in the large arteries, and—critically—do not account for cardiac output, thereby preventing application of their methods to pediatric patients with single ventricle heart disease and highly variable cardiac output.

By requiring a central (rather than peripheral) injection site, we reduce the likelihood of falsely accelerating the PCTT. And by not starting the transit time until contrast reaches the distal pulmonary vessels, we avoid most extra-parenchymal collateral vessels and focus analysis on the intra-parenchymal PAVMs. Indeed, in *ex vivo* studies of isolated, perfused lungs, the distal capillary bed was shown to be responsible for the major variations in pulmonary transit time (17).

While visual calculation of the PCTT is reliable and fast, we plan to develop an automated analysis for further ease of application. We anticipate that machine learning coupled with shadow tracking will prove effective against a background of respiratory and cardiac motion. Quantification of MR or CT-derived transit times will also be helpful, especially as their use increases for pre-Glenn evaluation (18, 19). Finally, as more data is gathered, we hope to use PCTT to create a continuous scale that could predict outcomes and pulmonary vascular complications after the Glenn and Fontan surgeries.

### Limitations

For clinical application, a direct comparison between our pulmonary capillary transit time and bubble contrast echocardiography will be necessary. Due to the variability of both anatomy and clinical management, collaboration with other centers will be necessary to power discovery of the patient-specific factors that increase risk of PAVM development.

## Conclusions

Pulmonary capillary transit time is a simple, rapid, and reproducible method to assess changes in the pulmonary vascular architecture. Our results confirm the traditional teaching that the contrast takes at least three cardiac cycles to traverse the normal pulmonary vasculature. Our longitudinal study of PCTT in single ventricle patients reinforces the hepatic factor hypothesis and shows a measurable effect of hepatic factor loss in even “normal” Glenn patients.

## Data Availability

All data produced in the present study are available upon request to the authors, pending successful approval of data use sharing agreement with the University of Texas Southwestern Medical Center.

## Acknowledgements

We would like the members of both the Cleaver Lab and the Leducq ReVAMP Network for their input and guidance.

## Ethical Standards

The authors assert that this work complies with the ethical standards of the relevant national guidelines and with the Helsinki Declaration of 1975, as revised in 2008, and has been approved by the University of Texas Southwestern Independent Review Board (IRB), UTSW IRB 2020-0047.

## REFERENCES

1. Cloutier A, Ash JM, Smallhorn JF, Williams WG, Trusler GA, Rowe RD, et al. Abnormal distribution of pulmonary blood flow after the Glenn shunt or Fontan procedure: risk of development of arteriovenous fistulae. Circulation. 1985;72(3):471–9. Epub 1985/09/01. doi: 10.1161/01.cir.72.3.471. PubMed PMID: 4017202.

2. McFaul RC, Tajik AJ, Mair DD, Danielson GK, Seward JB. Development of pulmonary arteriovenous shunt after superior vena cava-right pulmonary artery (Glenn) anastomosis. Report of four cases. Circulation. 1977;55(1):212–6. Epub 1977/01/01. doi: 10.1161/01.cir.55.1.212. PubMed PMID: 830212.

3. Srivastava D, Preminger T, Lock JE, Mandell V, Keane JF, Mayer JE, Jr., et al. Hepatic venous blood and the development of pulmonary arteriovenous malformations in congenital heart disease. Circulation. 1995;92(5):1217–22. Epub 1995/09/01. doi: 10.1161/01.cir.92.5.1217. PubMed PMID: 7648668.

4. Mathur M, Glenn WW. Long-term evaluation of cava-pulmonary artery anastomosis. Surgery. 1973;74(6):899–916. Epub 1973/12/01. PubMed PMID: 4127196.

5. Kim SJ, Bae EJ, Lee JY, Lim HG, Lee C, Lee CH. Inclusion of hepatic venous drainage in patients with pulmonary arteriovenous fistulas. Ann Thorac Surg. 2009;87(2):548–53. Epub 2009/01/24. doi: 10.1016/j.athoracsur.2008.10.024. PubMed PMID: 19161777.

6. McElhinney DB, Kreutzer J, Lang P, Mayer JE, Jr., del Nido PJ, Lock JE. Incorporation of the hepatic veins into the cavopulmonary circulation in patients with heterotaxy and pulmonary arteriovenous malformations after a Kawashima procedure. Ann Thorac Surg. 2005;80(5):1597–603. Epub 2005/10/26. doi: 10.1016/j.athoracsur.2005.05.101. PubMed PMID: 16242423.

7. Shah MJ, Rychik J, Fogel MA, Murphy JD, Jacobs ML. Pulmonary AV Malformations After Superior Cavopulmonary Connection: Resolution After Inclusion of Hepatic Veins in the Pulmonary Circulation. The Annals of Thoracic Surgery. 1997;63(4):960–3. doi: 10.1016/s0003-4975(96)00961-7.

8. Pike NA, Vricella LA, Feinstein JA, Black MD, Reitz BA. Regression of severe pulmonary arteriovenous malformations after Fontan revision and “hepatic factor” rerouting. Ann Thorac Surg. 2004;78(2):697–9. Epub 2004/07/28. doi: 10.1016/j.athoracsur.2004.02.003. PubMed PMID: 15276554.

9. McElhinney DB, Marx GR, Marshall AC, Mayer JE, Del Nido PJ. Cavopulmonary pathway modification in patients with heterotaxy and newly diagnosed or persistent pulmonary arteriovenous malformations after a modified Fontan operation. The Journal of Thoracic and Cardiovascular Surgery. 2011;141(6):1362–70.e1. doi: 10.1016/j.jtcvs.2010.08.088.

10. Spearman AD, Ginde S. Pulmonary Vascular Sequelae of Palliated Single Ventricle Circulation: Arteriovenous Malformations and Aortopulmonary Collaterals. Journal of Cardiovascular Development and Disease. 2022;9(9):309. doi: 10.3390/jcdd9090309.

11. Asada D, Morishita Y, Kawai Y, Kajiyama Y, Ikeda K. Efficacy of bubble contrast echocardiography in detecting pulmonary arteriovenous fistulas in children with univentricular heart after total cavopulmonary connection. Cardiol Young. 2020:1–4. Epub 2020/01/10. doi: 10.1017/S104795111900324X. PubMed PMID: 31916529.

12. Chang RK, Alejos JC, Atkinson D, Jensen R, Drant S, Galindo A, et al. Bubble contrast echocardiography in detecting pulmonary arteriovenous shunting in children with univentricular heart after cavopulmonary anastomosis. J Am Coll Cardiol. 1999;33(7):2052–8. Epub 1999/06/11. doi: 10.1016/s0735-1097(99)00096-0. PubMed PMID: 10362213.

13. Forde KA, Fallon MB, Krowka MJ, Sprys M, Goldberg DS, Krok KL, et al. Pulse Oximetry Is Insensitive for Detection of Hepatopulmonary Syndrome in Patients Evaluated for Liver Transplantation. Hepatology. 2019;69(1):270–81. doi: 10.1002/hep.30139.

14. Tonelli AR, Naal T, Dakkak W, Park MM, Dweik RA, Stoller JK. Assessing the kinetics of microbubble appearance in cirrhotic patients using transthoracic saline contrast-enhanced echocardiography. Echocardiography. 2017;34(10):1439–46. doi: 10.1111/echo.13662.

15. Faughnan ME, Mager JJ, Hetts SW, Palda VA, Lang-Robertson K, Buscarini E, et al. Second International Guidelines for the Diagnosis and Management of Hereditary Hemorrhagic Telangiectasia. Ann Intern Med. 2020;173(12):989–1001. Epub 2020/09/08. doi: 10.7326/M20-1443. PubMed PMID: 32894695.

16. Zhao H, Tsauo J, Zhang X, Ma H, Weng N, Wang L, et al. Pulmonary transit time derived from pulmonary angiography for the diagnosis of hepatopulmonary syndrome. Liver Int. 2018;38(11):1974–81. Epub 2018/03/25. doi: 10.1111/liv.13741. PubMed PMID: 29573542.

17. Clough AV, Haworth ST, Hanger CC, Wang J, Roerig DL, Linehan JH, et al. Transit time dispersion in the pulmonary arterial tree. J Appl Physiol (1985). 1998;85(2):565–74. Epub 1998/08/04. doi: 10.1152/jappl.1998.85.2.565. PubMed PMID: 9688734.

18. Brown DW, Gauvreau K, Powell AJ, Lang P, Colan SD, Del Nido PJ, et al. Cardiac magnetic resonance versus routine cardiac catheterization before bidirectional glenn anastomosis in infants with functional single ventricle: a prospective randomized trial. Circulation. 2007;116(23):2718–25. Epub 2007/11/21. doi: 10.1161/CIRCULATIONAHA.107.723213. PubMed PMID: 18025538.

19. James L, Tandon A, Nugent A, Malik S, Ramaciotti C, Greil G, et al. Rationalising the use of cardiac catheterisation before Glenn completion. Cardiol Young. 2018;28(5):719–24. Epub 2018/03/07. doi: 10.1017/S1047951118000240. PubMed PMID: 29506588.

