## Supplementary for "Angiographic Tool to Detect Pulmonary Arteriovenous Malformations in Single Ventricle Physiology"

**Table S1:**

| **Patient Group** | **Location of Catheter or Sheath** |
| --- | --- |
| Pulmonary Artery/Valve Stenosis | Main pulmonary artery or right ventricle |
| Pre-Glenn: Sano shunt | Primary ventricle |
| Pre-Glenn: mBTT shunt | Ipsilateral subclavian artery |
| Glenn | Superior caval vein |
| Fontan | Superior caval vein or Fontan conduit |

**Injection Site Locations:** The site of contrast injection was selected for each anatomical group. Angiograms with pulmonary angiograms in other locations were excluded from the study.

**Table S2:**

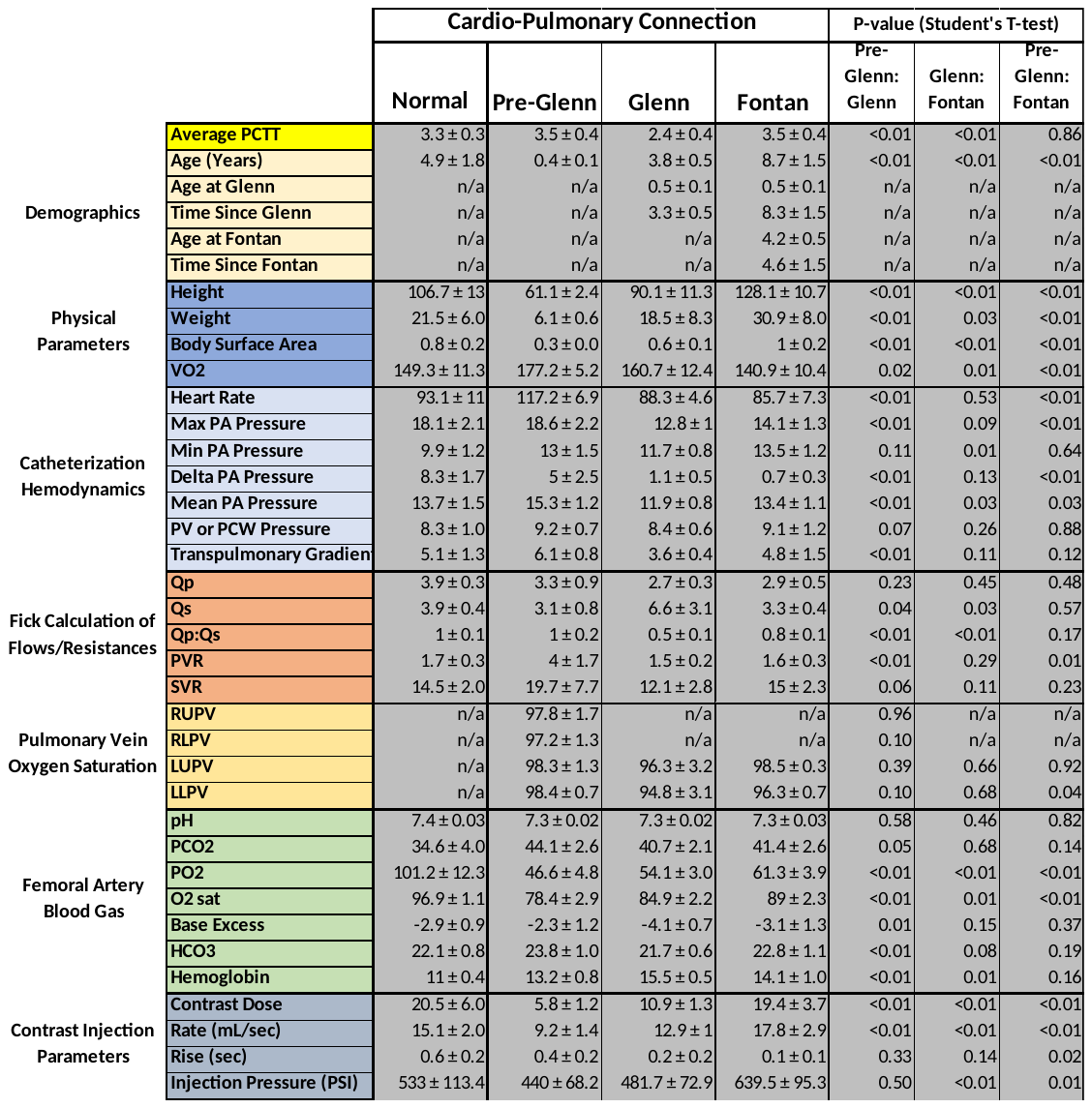

**Clinical Analysis of Longitudinal Cohort:** Clinical and catheterization data obtained at each surgical stage for the longitudinal cohort. Data is presented as mean ± value to obtain the 95% confidence interval.

**Supplementary Table S3:**

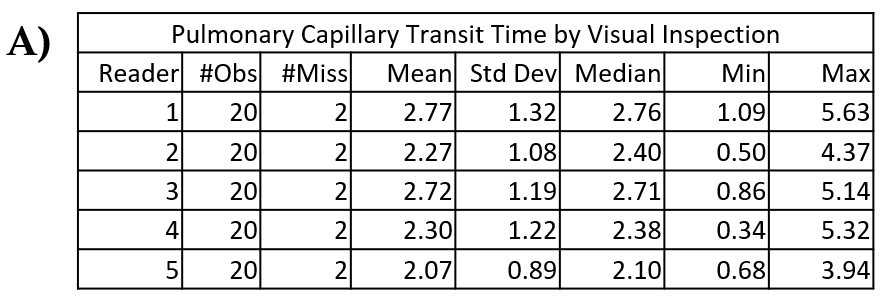

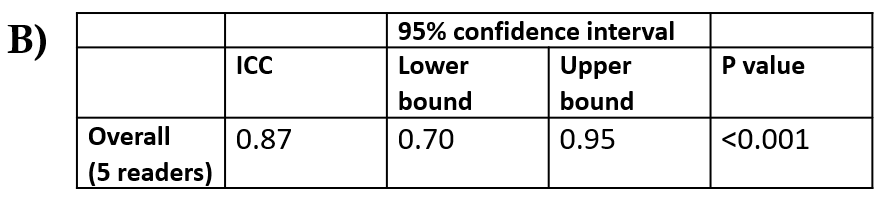

**Visual Calculation of PCTT is Reproducible Between Observers. (A)** Descriptive statistics for PCTT measurements obtained by five trained observers. **(B)** Intraclass correlation coefficient (ICC) based on the two-way random effects model was used to investigate the agreement between readers. The p values for pairwise ICC were not adjusted.

**Figure S1:**

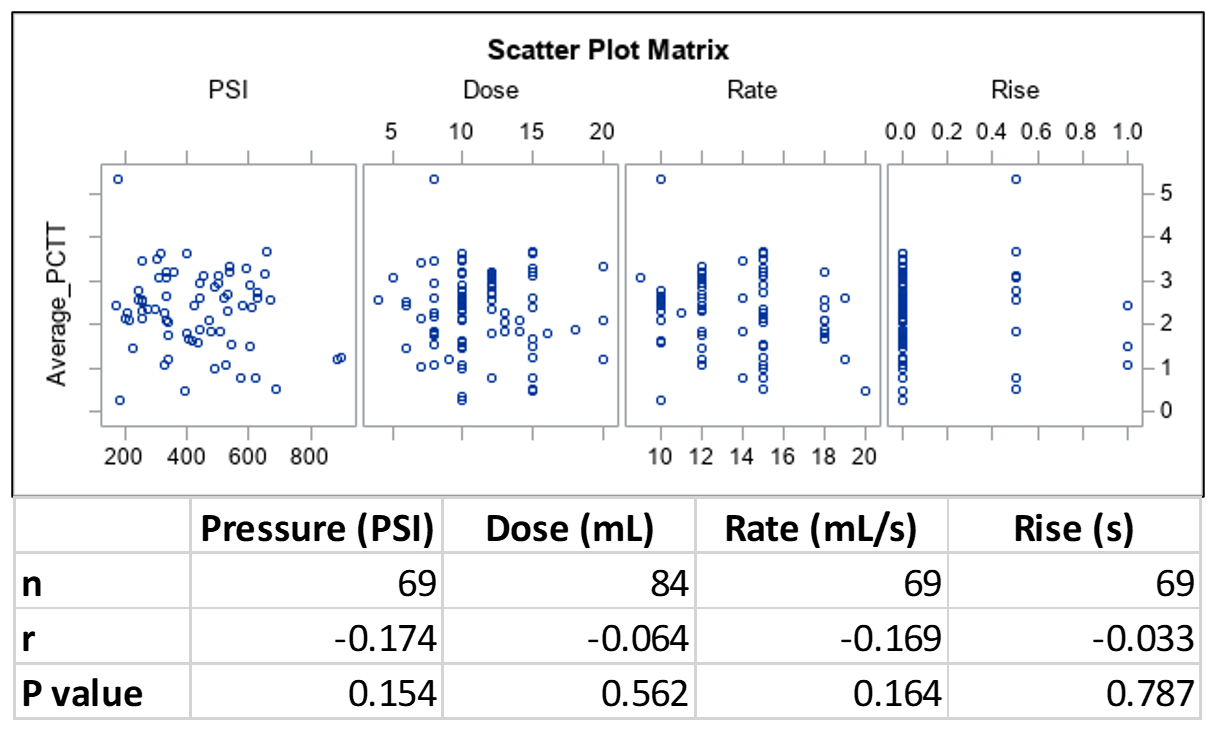

**PCTT Does not Correlate with Injection Parameters in Glenn Patients.** Scatterplot shows lack of correlation of PCTT with injection pressure (PSI), dose (mL), rate (mL/s), or rate of rise (s).

**Figure S2:**

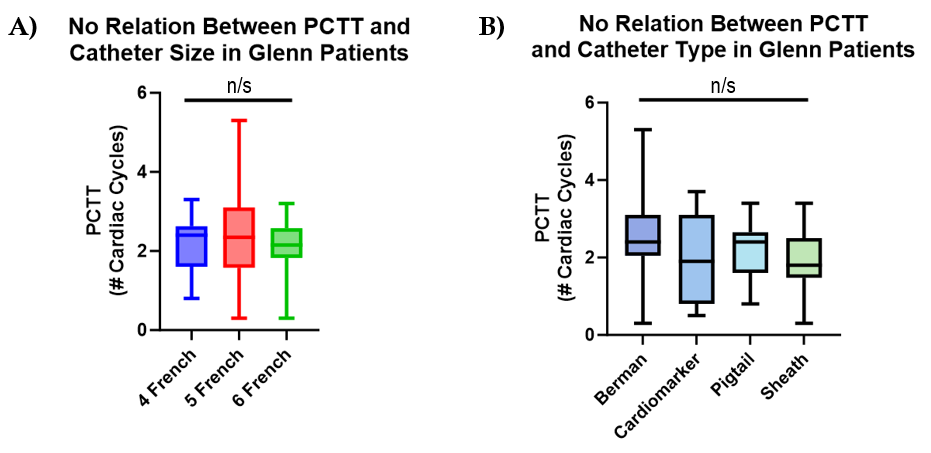

**Relation of Catheter Size and Type to PCTT. (A)** Catheter size does not significantly impact mean PCTT. **(B)** Catheter type does not significantly impact PCTT.
